# Hematoma Evacuation and Risk of Subsequent Ischemic Stroke and Coronary Ischemic Events: MISTIE III and ATACH-2 Analysis

**DOI:** 10.1101/2025.11.20.25340710

**Authors:** Wendy C. Ziai, William Harris, Adnan I. Qureshi, Issam Awad, Daniel F. Hanley, Santosh B. Murthy

## Abstract

**BACKGROUND:** Nontraumatic intracerebral hemorrhage (ICH) especially in deep locations is independently associated with a long-term increased risk of major arterial ischemic events. Minimally invasive surgery (MIS) has differential impact on outcomes by location. Whether ischemic events modify outcomes after MIS and the influence of ICH location is poorly understood.

**METHODS:** We pooled individual patient data from the MISTIE III and ATACH-2 trials. The exposure was ICH location (deep vs. lobar). The outcome was a symptomatic, clinically overt ischemic stroke or coronary ischemic event. We evaluated the association between ICH location and risk of an ischemic event using Cox regression analyses after adjustment for demographics, vascular comorbidities, and ICH characteristics. We investigated whether ischemic events modified the impact of effective MIS, defined as end of treatment volume (EOT) <15 mL, on modified Rankin scale (mRS) 0-3 at one year in MISTIE III using logistic regression.

**RESULTS:** Of 1470 ICH patients median hematoma volume was 17.3 mL (interquartile range, 7.2–35.7) and 1186 (80.7%) were deep. During a median follow-up of 110 days (iqr 110-365), 70 ischemic events occurred, 60 (5.0% cumulative incidence) in patients with deep ICH and 10 after lobar ICH (cumulative incidence 3.2%). In adjusted analyses, deep ICH location was associated with an increased risk of ischemic events (hazard ratio, 2.3 [95% CI, 1.1–4.8]), but MIS was not. In the full MISTIE cohort, in patients without ischemic events during follow-up, MIS with EOT<15 mL was significantly associated with favorable one year outcome (OR 1.90 (95% CI: 1.16-3.12; P for interaction = 0.04). There was no effect modification for deep location (P for interaction = 0.128). For lobar ICH, EOT ICH volume <15 mL with MIS was associated with good outcome regardless of ischemic events.

**CONCLUSIONS:** In a heterogeneous cohort of patients with ICH, deep ICH location was associated with increased risk of ischemic events over the short term, but this appears to have low impact on one-year outcomes with successful surgery.

## Introduction

Nontraumatic intracerebral hemorrhage (ICH) is responsible for high mortality and stroke-related disability. (1) ICH survivors are at high risk of complications including major vascular events; these include ICH recurrence with an annual risk of 1.3% to 7.4% (2) and ischemic arterial events such as ischemic stroke and myocardial infarction (MI), which are at least as frequent (3). ICH location is an important factor when evaluating these risks because lobar ICH is etiologically related to cerebral amyloid angiopathy and is associated with significantly higher ICH recurrence rates than deep ICH, which is caused by deep perforator vasculopathy and tends to be associated with significantly higher future ischemic events (4,5). We have previously shown that among patients with ICH, a new ischemic arterial event results in 50% lower odds of favorable one year outcome (6) and 2-fold higher risk of mortality (7).

Minimally invasive surgery (MIS) for supratentorial ICH has begun to show significant benefits especially when hematoma reduction goals are met (8). The Early Minimally Invasive Removal of ICH (ENRICH) clinical trial found that early MIS can improve functional outcomes, particularly for those with lobar hemorrhages (9). Deep ICH did not seem to benefit from MIS in ENRICH and in the MIS Plus Alteplase in ICH Evacuation (MISTIE III) clinical trial, where 61% of the population had deep hemorrhages, patients with deep ICH had worse outcomes despite having smaller hematoma volumes and greater volume reduction with MIS (8,9).

It is unclear whether the relatively worse outcomes after MIS for deep ICH are secondary to the hematoma location near deep structures which lead to higher NIHSS and lower GCS, to tissue injury from surgery, or to medical complications, especially arterial ischemic events that occur over the longer term. Better knowledge of these relationships will better inform the potential benefit of surgery, especially for deep ICH. We therefore sought to evaluate the association between MIS and the subsequent risk of an arterial ischemic event compared to medical management alone, whether this risk differed by ICH location, and whether arterial ischemic events modified one-year outcomes after effective MIS. We assessed this relationship in two large, U.S. based clinical trials comprising a diverse cohort of patients with ICH, treated with MIS or medical management alone.

## Methods

### Data Availability

The MISTIE III (8) and the ATACH-2 (Antihypertensive Treatment of Acute Cerebral Hemorrhage-2) (10) trial data, including de-identified participant data, is available at the National Institute of Neurological Disorders and Stroke data archive (https://www.ninds.nih.gov/Current-Research/Research-Funded-NINDS/Clinical-Research/Archived-Clinical-ResearchDatasets). Those seeking access must submit a NINDS data request proposal and receive approval.

### Study Design

This is a retrospective cohort study using pooled individual-level data from ICH patients enrolled in 2 clinical trials, the MISTIE III trial (8) and the ATACH-2 trial (10). Trial protocols were approved by the ethics committee at each study site and written informed consent for research was obtained from all participants (or legal representatives or surrogates when applicable). The Johns Hopkins Hospital institutional review board approved the de-identified analysis of previously collected patient data. This study was performed in accordance with the Strengthening the Reporting of Observational Studies in Epidemiology guidelines for reporting observational studies. (11)

### Study Setting

MISTIE III was a randomized, controlled, open-label, phase 3 trial that evaluated minimally invasive surgery (MIS) with stereotactic thrombolysis (n=250) compared to standard medical care (n=249). (8) Patients had moderate to large non-traumatic, supratentorial ICH. The trial was neutral on the primary endpoint of modified Rankin scale score (mRS) 0-3 at one year although there was a mortality benefit with MIS and on secondary analysis, a functional outcome benefit for surgical subjects who achieved the surgical goal of <15 mL ICH volume at the end of treatment (EOT). Enrollment criteria included age ≥18 years, ICH ≥30 mL, Glasgow Coma Scale score (GCS) of ≤14 or National Institutes of Health Stroke Scale score ≥6, premorbid mRS 0 or 1, and radiographic ICH volume stability for at least 6 hours.

ATACH-2 was a randomized, multicenter, open-label trial that compared the efficacy of intensive versus standard blood pressure management starting within 4.5 hours of ICH onset in patients with spontaneous supratentorial ICH. (10) There was no difference in the primary end point of mRS 4-6 at 90 days. The enrollment criteria for ATACH-2 were age ≥18 years, presentation GCS of ≥5 at presentation and initial hematoma volume <60 mL. Both trials excluded secondary causes of ICH and patients with planned withdrawal of life sustaining treatment.

### Measurements

We included all patients with ICH enrolled in the MISTIE III and ATACH-2 trials. The exposure was ICH location (deep vs. lobar) ascertained on the computed tomography scan at the time of admission. Lobar hematoma location was defined by selective involvement of cerebral cortex, underlying white matter, or both, and deep ICH location by selective involvement of thalami, basal ganglia, or both. In each study, a neuroimaging core of board-certified neurologists and radiologists blinded to treatment assignment and outcome evaluated the computed tomography scans of the head. Magnetic resonance imaging (MRI) scans were obtained at the discretion of the treating physicians in the ATACH-2 trial.(12) In the MISTIE III trial, 1 of 2 protocolized MRI scans was obtained during hospitalization—on the day of randomization and between days 7 and 10 after randomization. Exceptions included unstable patients, withdrawal of life-sustaining treatment, or contraindications for MRI.(13)

The outcome was a symptomatic, clinically overt ischemic stroke or coronary ischemic event, adjudicated centrally within each trial as previously defined.(12,13) Clinical criteria for ischemic stroke included focal or global neurological deficits (motor, sensory, cranial nerve or speech). Radiological criteria on either CT or MRI required presence of focal and/or global abnormalities indicating areas of lost tissue viability. Only symptomatic events were included in this study.

Clinical criteria for coronary ischemic events included acute coronary syndrome, congestive heart failure exacerbation, and cardiac arrest. Acute coronary syndrome was defined per the 2018 consensus criteria (14) Mild troponin elevations in the first week after ICH which typically indicate acute stress cardiomyopathy were excluded.

All ischemic strokes and coronary ischemic events were reported by site investigators, independently reviewed by each trial coordinating center and adjudicated by a safety events committee. Follow-up data for MISTIE III was accrued at 3 months, 6 months and one year while ATACH-2 only had 90-day outcomes. To maintain uniform follow-up times, in the outcome analysis we only counted the first 120 days after discharge, including a 30-day grace period for ascertaining 3-month outcomes.

### Statistical Analysis

We summarized normally distributed continuous variables as means with standard deviations, while non-normally distributed variables were reported as medians with interquartile range (IQR). For univariate analyses, we used Wilcoxon rank sum test or Student’s t-test for continuous variables depending on the normality of distribution, and Chi-squared test or Fisher’s exact test for categorical variables. Kaplan-Meier survival statistics were used to estimate the incidence of stroke or coronary ischemic events among participants based on the median hematoma location.

In the primary analyses, we evaluated the association between ICH location and risk of an ischemic event using Cox regression analyses after adjustment for demographics, vascular comorbidities, and ICH characteristics. Since there were a total of 66 ischemic events, we built 4 different models adjusting for 4-6 covariates at a time, chosen apriori to prevent overfitting of the model. The first model was unadjusted; the second was adjusted for universal confounders, age, sex and race; the third included age and vascular comorbidities (atrial fibrillation, prior stroke, or transient ischemic attack); fourth included age and diabetes mellitus, hyperlipidemia and atrial fibrillation; the fifth included ICH characteristics (age, presence of intraventricular hemorrhage [IVH], treatment randomization arm). All models were adjusted for baseline ICH volume.

Patients were censored at the time of death, occurrence of outcome, or at the last available follow-up (day 110). We assessed the overall fit of the model using Cox-Snell residuals and constructed log-log plots to ensure that the proportional hazard assumption was not violated. In secondary analyses, we excluded patients randomized to the intervention arms of MISTIE III and ATACH-2.

### Effect modification

We used logistic regression to assess whether ischemic events modified the effect of MIS with end of treatment volume (EOT) <15mL on modified Rankin scale (mRS) 0-3 at one year in MISTIE III. All sensitivity analyses used the 5 Cox regression models described above. Statistical analyses were performed using Stata (StataCorp, Statistical Software: Release 16, College Station, TX; StataCorp LP; 2019). All analyses were 2 tailed and the threshold for statistical significance was P<0.05.

## Results

Of 1499 patients with ICH, 15 were excluded who were lost to follow-up and 2 had missing data on ICH location (Figure 1). Of 1482 patients included in the analysis, the mean age was 61.7 (SD, 12.8) years, and 569 (38.4%) were female. Baseline characteristics of patients by study are shown in Table 1 and Tables S1 and S2 and by ICH location in Table 2. The median hematoma volume was 17.4 mL (interquartile range, 7.2-35.7). When stratified by deep versus lobar location, patients with deep ICH were younger, less likely to be female, with fewer cardiovascular risk factors, and higher severity of ICH at presentation.

**Figure 1:**
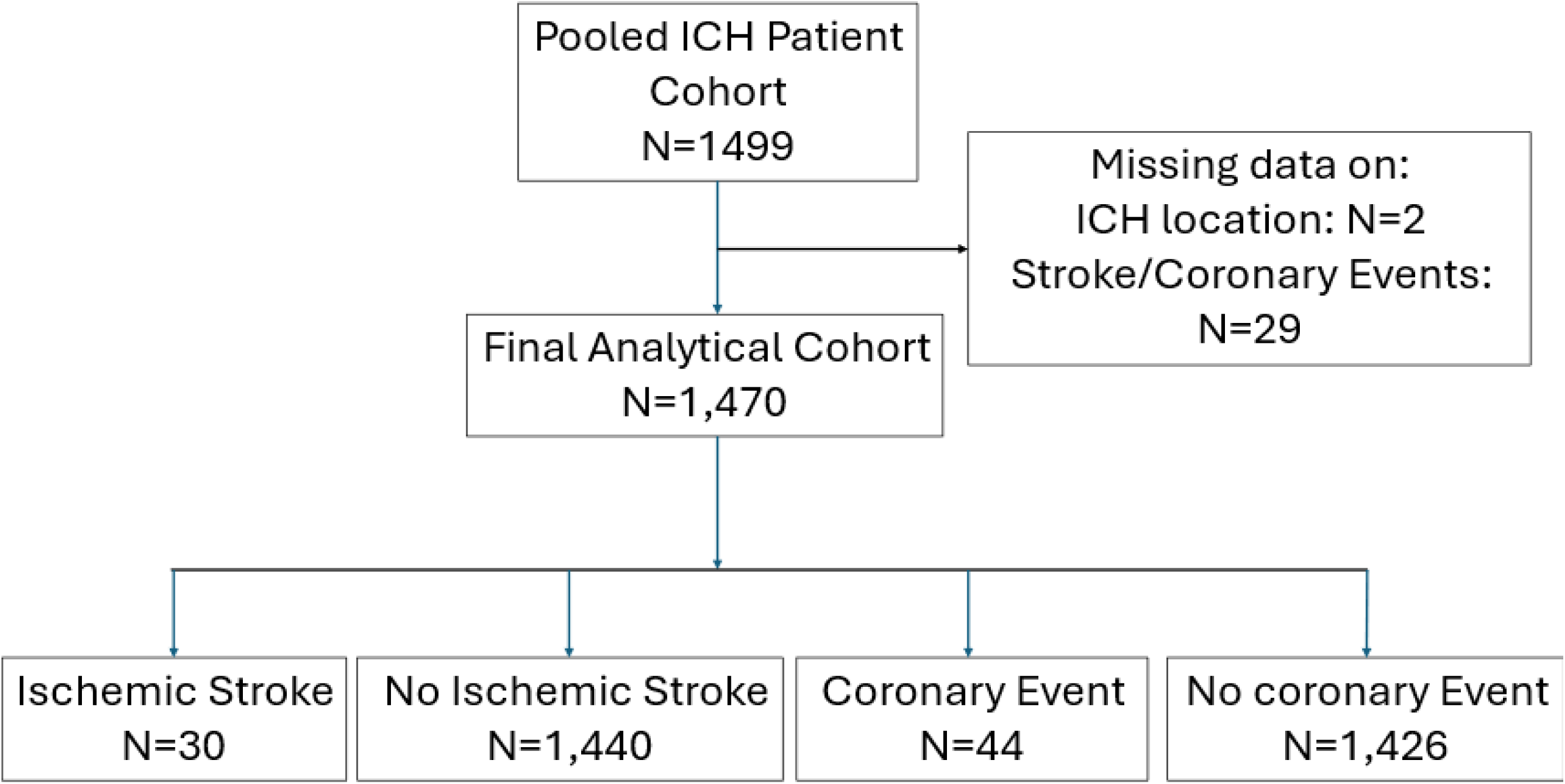
Flowchart Showing Patient Selection Abbreviations: ICH, intracerebral hemorrhage.

**Table 1.**
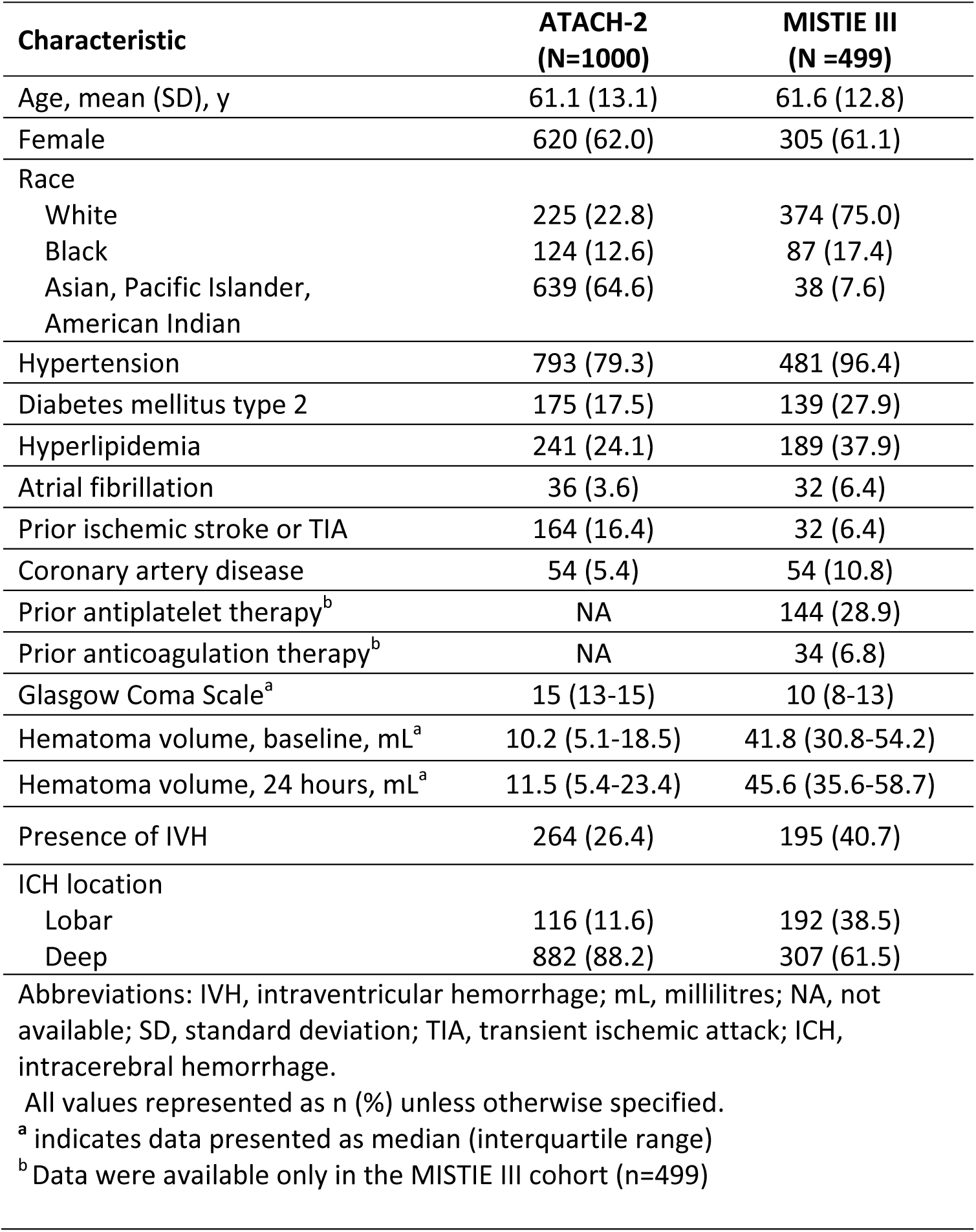
Baseline Characteristics of ICH Patients, Stratified by Study Cohort.

| Characteristic | ATACH-2<br>(N=1000) | MISTIE III<br>(N =499) |
| --- | --- | --- |
| Age, mean (SD), y | 61.1 (13.1) | 61.6 (12.8) |
| Female | 620 (62.0) | 305 (61.1) |
| Race |  |  |
| White | 225 (22.8) | 374 (75.0) |
| Black | 124 (12.6) | 87 (17.4) |
| Asian, Pacific Islander,<br>American Indian | 639 (64.6) | 38 (7.6) |
| Hypertension | 793 (79.3) | 481 (96.4) |
| Diabetes mellitus type 2 | 175 (17.5) | 139 (27.9) |
| Hyperlipidemia | 241 (24.1) | 189 (37.9) |
| Atrial fibrillation | 36 (3.6) | 32 (6.4) |
| Prior ischemic stroke or TIA | 164 (16.4) | 32 (6.4) |
| Coronary artery disease | 54 (5.4) | 54 (10.8) |
| Prior antiplatelet therapy<sup>b</sup> | NA | 144 (28.9) |
| Prior anticoagulation therapy<sup>b</sup> | NA | 34 (6.8) |
| Glasgow Coma Scale<sup>a</sup> | 15 (13-15) | 10 (8-13) |
| Hematoma volume, baseline, mL<sup>a</sup> | 10.2 (5.1-18.5) | 41.8 (30.8-54.2) |
| Hematoma volume, 24 hours, mL<sup>a</sup> | 11.5 (5.4-23.4) | 45.6 (35.6-58.7) |
| Presence of IVH | 264 (26.4) | 195 (40.7) |
| ICH location |  |  |
| Lobar | 116 (11.6) | 192 (38.5) |
| Deep | 882 (88.2) | 307 (61.5) |

Abbreviations: IVH, intraventricular hemorrhage; mL, millilitres; NA, not available; SD, standard deviation; TIA, transient ischemic attack; ICH, intracerebral hemorrhage.
All values represented as n (%) unless otherwise specified.
<sup>a</sup> indicates data presented as median (interquartile range)
<sup>b</sup> Data were available only in the MISTIE III cohort (n=499)

**Table 2.** Baseline Characteristics of ICH Patients, Stratified by ICH Location.

| Characteristic | Deep<br>(N=1179) | Lobar<br>(N =303) | P value |
| --- | --- | --- | --- |
| Age, mean (SD), y | 60.4 (12.9) | 66.8 (11.3) | <0.001 |
| Female | 427 (36.2) | 142 (46.9) | 0.001 |
| Race+ |  |  | <0.001 |
| White | 394 (33.7) | 197 (65.5) |  |
| Black | 155 (13.3) | 51 (16.9) |  |
| Asian, Pacific Islander,<br>American Indian | 620 (53.0) | 53 (17.6) |  |
| Hypertension | 989 (85.7) | 271 (90.0) | 0.09 |
| Diabetes mellitus type 2 | 239 (20.5) | 74 (24.7) | 0.14 |
| Hyperlipidemia | 289 (25.6) | 140 (47.6) | <0.001 |
| Atrial fibrillation | 40 (3.4) | 27 (8.9) | <0.001 |
| Prior ischemic stroke or TIA | 164 (14) | 30 (9.9) | 0.05 |
| Coronary artery disease | 66 (5.6) | 41 (13.5) | <0.001 |
| Systolic blood pressure | 196 (180-217) | 181 (160-204) | <0.001 |
| Glasgow Coma Scale<sup>a</sup> | 14 (11-15) | 13 (10-15) | <0.001 |
| Hematoma volume, baseline, mL<sup>a</sup> | 14.9 (6.6-30.3) | 34.1 (11.9-52.1) | <0.001 |
| Hematoma volume, 24 hours, mL<sup>a</sup> | 11.5 (5.4-23.4) | 45.6 (35.6-58.7) |  |
| Presence of IVH | 360 (31.1) | 98 (33.3) | 0.45 |
| Treatment with MIS | 163 (13.8) | 87 (28.7) | <0.001 |

Abbreviations: ICH, intracerebral hemorrhage; IVH, intraventricular hemorrhage; mL, millilitres; SD, standard deviation; TIA, transient ischemic attack; MIS, minimally invasive surgery. All values represented as n (%) unless otherwise specified. <sup>a</sup> indicates data presented as median (interquartile range)

### Primary Analysis

During a median of 110 days (IQR 110-365), a total of 67 arterial ischemic events (30 ischemic strokes and 41 cardiac events; 4 patients with one of each) occurred of which 58 were in patients with deep ICH (4.9% cumulative incidence) and 9 were in patients with lobar ICH (cumulative incidence 2.9%). Table 3 shows multivariate Cox regression models using hematoma location dichotomized as deep vs. lobar. The first model was unadjusted and did not show a significant association of deep ICH with risk of an arterial ischemic event (Table 3) (hazard ratio [HR], 1.6 [95% confidence interval [CI] 0.8-3.3]). In all 4 adjusted Cox regression models, deep ICH location was associated with an increased risk of an arterial ischemic event; in model 2 after adjusting for demographics (HR, 2.2 [95% CI, 1.06-4.8]); in model 3 after accounting for age, CAD, HTN and prior stroke or transient ischemic attack (HR, 2.4 [95% CI, 1.1-5.1]); in model 4 controlling for age, atrial fibrillation, hyperlipidemia and diabetes mellitus (HR, 2.3 [95% CI, 1.1-4.9]); and finally, in model 4 that controlled for age, the presence of IVH, and treatment with MIS (HR, 2.3 [95% CI, 1.1–4.8].

**Table 3.** Cox Regression Analyses Showing the Relationship Between Hematoma Location and Risk of a Cardiac Ischemic Event or Ischemic Stroke Among ICH Survivors.

| Cox Regression Models | Stratified by ICH Location (Deep vs. Lobar) |  |
| --- | --- | --- |
|  | HR (95% CI) | p value |
| Unadjusted | 1.6 (0.8-3.3) | 0.18 |
| **Model 1:** Adjusted for age, sex, race, ICH volume | 2.2 (1.1-4.8) | 0.03 |
| **Model 2:** Adjusted for age, vascular comorbidities (CAD, HTN, prior stroke or TIA) and ICH volume | 2.4 (1.1-5.1) | 0.02 |
| **Model 3:** Adjusted for age, vascular comorbidities (atrial fibrillation, hyperlipidemia, DM) and ICH volume | 2.3 (1.1-4.9) | 0.03 |
| **Model 4:** Adjusted for ICH characteristics (age, ICH volume, presence of IVH, treatment with MIS) | 2.3 (1.1-4.8) | 0.03 |
| Abbreviations: ICH, intracerebral hemorrhage; HR, hazard ratio; CI, confidence interval; CAD, coronary artery disease; HTN, hypertension; DM, diabetes mellitus; TIA, transient ischemic attack; IVH, intraventricular hemorrhage; MIS, minimally invasive surgery. |  |  |

### Secondary analysis

In sensitivity analyses, after excluding 250 patients randomized to surgical hematoma evacuation in the MISTIE III trial and after the exclusion of 500 patients in the ATACH-2 trial who received intensive blood pressure reduction, we no longer observed a significant association between deep ICH location and an increased risk of arterial ischemic events. These results may reflect small numbers of events in subgroups from the trials (Table S1).

#### Effect modification

In the logistic regression model for one year outcome in MISTIE 3 patients only, adjusting for age, sex, enrollment GCS, baseline ICH volume, deep ICH location, and presence of IVH, the interaction of EOT volume < 15mL with MACE was significant. Odds of a good functional outcome were significantly greater in absence of MACE Adjusted OR [95% CI] 1.68 [1.03 – 2.76] compared to presence of MACE (aOR [95% CI] 0.15 [0.01 – 1.53]; P _interaction_ = 0.038) (Figure 2).

**Figure 2:**
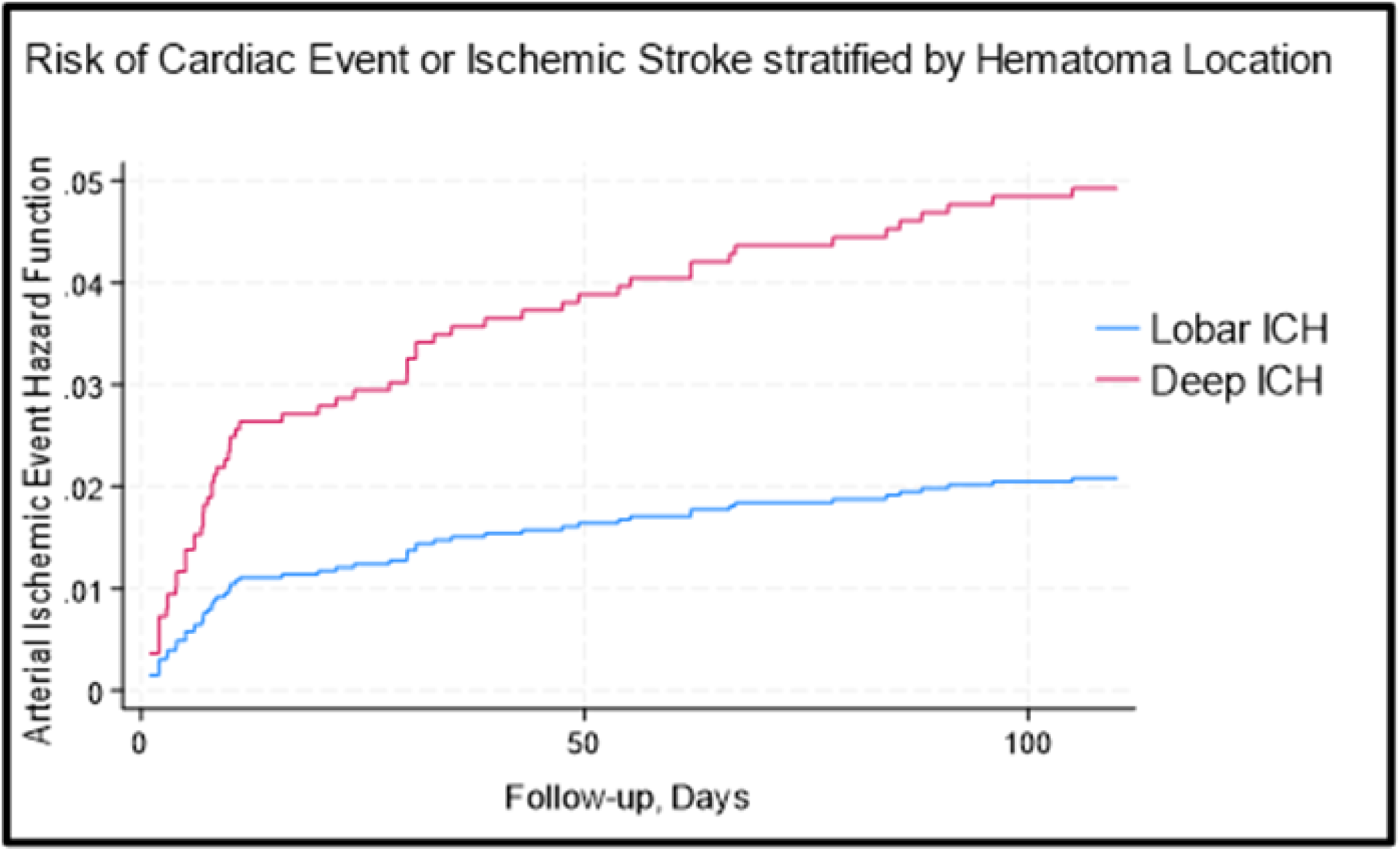
Kaplan-Meier Survival Curve Showing Risk of a Cardiac Ischemic Event or Ischemic Stroke, Stratified by Deep and Lobar Hematoma Location. Abbreviations: ICH, intracerebral hemorrhage.

## Discussion

In a pooled heterogeneous clinical trial cohort of patients with ICH, deep ICH location was independently associated with an increased short-term risk of an arterial ischemic event in adjusted analyses, but not without adjustment for risk factors. The risk of either an ischemic stroke or cardiac ischemic event increased by about 2% for deep hematomas compared with lobar ICH. In MISTIE III patients, absence of an ischemic event modified one-year functional outcome when achieving successful hematoma removal which was not, however, dependent on hematoma location.

Recent studies have shown a significantly increased risk of an arterial ischemic event in the first 6 months after ICH(15). Among U.S. Medicare beneficiaries, the 1-year cumulative incidence of an arterial ischemic event was 5.7% (95% CI, 4.8–6.8) in patients with ICH, which is consistent with our 3-month incidence of 4.5%. The distribution of these events, however, differs from our data which shows more cardiac ischemic events compared to ischemic strokes. In the Medicare beneficiary study, the risk of acute ischemic stroke was significantly increased during the first 6 months after ICH, but the risk of MI was not. Due to the retrospective nature of that study using claims data, it is possible that covert coronary ischemic events were missed leading to their underestimation.

The risk of arterial ischemic events seems to be associated more strongly with deep hematoma location.(16,17) Our findings align with that from a prospective observational cohort with 5-year follow-up where deep ICH was associated with a 6 times higher incidence of ischemic events than the incidence of hemorrhagic events which were more frequent in lobar ICH patients. In this cohort with a majority of deep ICH, ischemic stroke was still more common, however, compared to cardiac ischemic events. The use of protocolized adjudication of all ischemic events in the clinical trials from our study may explain the higher frequency of cardiac events in the short term. Also, we included cardiac arrest, acute coronary syndrome and congestive heart failure exacerbation in our definition of cardiac events, similar to that in cardiovascular trials, while these other studies only included myocardial infarction.

Several hypotheses have been proposed to explain the increased risk of arterial ischemic events in the first months after ICH. The cessation of antithrombotic drugs after ICH and unclear time for resumption especially in the first few months may have been a factor with approximately 36% of MISTIE III patients being on antithrombotics or anticoagulants prior to ICH onset. In the E-START trial of early versus late initiation of antiplatelet therapy after surgery for predominantly deep ICH (63%), starting acetylsalicylic acid on the third day after surgery in Chinese patients at high risk of ischemic events resulted in fewer postoperative ischemic major cardiovascular, cerebrovascular, or peripheral vascular events at 90 days compared to starting therapy at 30 days.(18) In the COCROACH prospective meta-analysis of clinical trials which included participants with predominantly non-lobar (63%) ICH and atrial fibrillation, resumption of oral anticoagulation reduced the risk of ischemic major adverse cardiovascular events at 2-6 years of follow-up compared to avoidance of anticoagulation (or use of antiplatelet agents) (19). These data contribute to the knowledge about heightened risk of ischemic events after ICH, and the need for strategies for improving long-term outcomes for non-lobar ICH.

Finally, severity of underlying cerebral small vessel disease, larger ICH volume, and activation of thrombo-inflammatory cascades by the hematoma products, both centrally and peripherally have all been associated with increased risk of ischemic events after ICH (20-22) and are supportive of our finding that effect modification by arterial ischemic events on outcomes in patients treated with MIS occurs without regard to hematoma location. In these severe ICH patients, worse comorbidity management may also be a factor.(23) From a cross-sectional study of patients with ICH in the Get With The Guidelines-Stroke registry, between 2011 and 2021, during the time of these trials, relatively few patients with ICH were prescribed antithrombotic or statin therapies at hospital discharge.(24)

Several limitations of this study are note-worthy. Data from our clinical trials may not be generalizable to the ICH population at large. Second, although we suspect low use of antithrombotic medications post ICH, data on secondary stroke prevention management was not available. Third, we were not able to assess the severity of pre-existing small vessel disease as only a subset of patients had magnetic resonance imaging of the brain. Fourth, although the number of cardiac and cerebral ischemic events was low which limited the number of covariates in the Cox regression models, the HR was similarly elevated across models with different combinations of covariates. Finally, although the vast majority of these 2 cohorts likely had hypertensive arteriolosclerosis as the etiology of ICH, we did not have more specific data on ICH etiology or cardiac risk factors which may have impacted on the risk of recurrent ischemic events.

## Conclusions

In a large prospective cohort of ICH survivors, deep hemorrhage location was associated with an increased short-term risk of major arterial ischemic events. In patients with successful MIS, absence of ischemic events positively affected functional recovery after ICH, regardless of hematoma location. Further study on strategies for improving secondary prevention of ischemic events after ICH is warranted.

## Nonstandard Abbreviations and Acronyms

ATACH-2: Antihypertensive Treatment of Acute Cerebral Hemorrhage-2 (ATACH-2) trial
EOT: End of treatment
ICH: Intracerebral hemorrhage
IVH: Intraventricular hemorrhage
MACCE: Major adverse cardiovascular and cerebrovascular events
MISTIE III: Minimally Invasive Surgery Plus Alteplase for Intracerebral Hemorrhage Evacuation Phase 3 trial

## Acknowledgments

We thank the patients involved in the ATACH-2 and MISTIE III trials, their families, and the investigators and coordinators who cared for them.

## Author Contributions

Dr. Ziai had full access to all of the data in the study and takes responsibility for the integrity of the data and the accuracy of the data analysis.

*Study concept and design*: Ziai, Murthy

*Acquisition, analysis, or interpretation of data*: Ziai, Harris, Murthy

*Drafting of the manuscript*: Ziai

*Critical revision of the manuscript for important intellectual content:* Ziai, Harris, Awad, Hanley, Murthy.

*Statistical analysis:* Ziai.

*Administrative, technical, or material support:* Ziai, Hanley.

*Study supervision:* Ziai.

